# Silicosis Prevalence and Associated Occupational Risk Factors Among Cassiterite (Tin Ore) Miners in Eastern Rwanda: A Cross-Sectional Analysis of Mining Practice and Risk in an Active and Retired Mining Cohort

**DOI:** 10.1101/2025.01.17.24319661

**Authors:** Samuel Hatfield, Symaque Dusabeyezu, Alphonse Nshimyiryo, Anne Niyigena, Peter Barebwanuwe, Michael Miller, Robert Tumusime, Jean Nepomuscene Renzaho, Michel Niyonsenga, Leah Traube, Tarek Elkady, Daniel Mays, Wellars Dusingizimana, Stella Savarimuthu, Paul D. Sonenthal, Phoebe Mwiseneza, Joel M Mubiligi, Fredrick Kateera, Innocent Kamali, Vincent K Cubaka

**Author notes:** **Corresponding Author:** Samuel Hatfield.

## Abstract

**Background:** Silicosis is one of the most common forms of pneumoconiosis worldwide. In Rwanda - there is paucity of data on the silicosis burden and occupational risk among underground miners.

**Methods:** We conducted a cross-sectional study among all miners from 8 cassiterite (tin ore) mining sites in Kayonza district, Eastern Rwanda. Questionnaire data and chest radiography were collected at Rwinkwavu District Hospital. Two radiologists reviewed all the chest radiography using ILO criteria, with a third radiologist reviewing films with ILO rating discrepancies and serving as an arbiter. Logistic regression was performed to investigate risk factors associated with radiographic silicosis.

**Findings:** In total, 1,021 mine workers were included in the primary outcome (risk) analysis. The median age was 32 years (IQR 26-40), and 948 participants (93%) were male. Of all participants, 94 (9%) were diagnosed with silicosis. Increased odds of silicosis were associated with working in a blasting station (aOR 3·30; 95% CI 1·68-6·45), excavation station (aOR 2·77; 95% CI 1·09-7·04), drilling station (aOR 2·51; 95% CI 1· 34-4· 70), exposure to tobacco (aOR 1·92; 95% CI 1· 14-3· 24), and increased time of working in mining (aOR 1·05; 95% CI 1·01-1·09).

**Interpretation:** Almost 1 in 10 Cassiterite miners in an occupational cohort of active and retired miners in the Eastern Province of Rwanda were found to have Silicosis. Tobacco use, certain mining tasks and duration of mining employment were significantly associated with increased risk of having a silicosis diagnosis. Our results indicate that screening and preliminary occupational risk analysis in a rural mining cohort is technically feasible.

**Funding:** All funding was provided by internal grants through the Non-Communicable Diseases program through Partners in Health/Inshuti Mu Buzima

**Research In Context:** *What is Already Known on this Topic:* Silicosis is recognized as a “Disease of Concern” for Low- and Middle-Income countries (LMICs) and is resurgent in parts of the United States and Australia. While the pathophysiology and preventative interventions necessary to address the development of silicosis, Little is known about the risk of mining related lung disease in Eastern Africa and no studies have been done in cassiterite miners in areas under conflict minerals regulation in our review of the literature.

*What this Study Adds:* The objectives of this study were threefold. First, to determine the radiographic prevalence of silicosis in a mining cohort in Eastern Rwanda. Second, to identify risk factors associated with the development of silicosis present in cassiterite mining in semi-formal mines. Third, to develop a methodology for identifying and tracking mining-related disease in a rural community reliant on the mining economy.

*How this study might affect research, practice or policy:* Our paper, in combination with available evidence, supports that silicosis continues to be a significant cause of morbidity and mortality in underground mining populations. It elucidates mechanisms within mining practice in Eastern Africa likely contributing to a high rate of advanced and accelerated silicosis in a region typically difficult to access for clinical surveillance programs. It also adds evidence of occupational risk associated with a pervasive and common mineral, cassiterite. Finally, it calls into question epidemiologic data on the true rates of silicosis in mining communities in Eastern Africa.

## Introduction

Silicosis, a form of occupational pneumoconiosis, is a chronic, incurable lung disease caused by inhaling silica dust leading to lung inflammation and the formation of irreversible fibrosis.^1^ Respirable crystalline silica (RCS) consists of tiny particles less than 10 micrometres in diameter and is commonly generated, or entrained into the air, during mining operations and in a variety of industries including construction work, quarrying, stone cutting, and hydraulic fracturing.^2^ Underground mining, with techniques such as those used in cassiterite mining in Eastern Rwanda, has a well-established association with silicosis.^3^

The current global prevalence of silicosis is not well defined, due to limited and/or absent screening programs in many low- and middle-income countries (LMICs). Silicosis is increasingly recognized as a pneumoconiosis of concern in LMICs, with trends showing increasing incidence in these countries compared to relative improvement in high-income nations. ^4^ Silicosis has been on the agendas of the International Labour Organization (ILO) and World Health Organization (WHO) and remains an urgent public health issue.^5^

Today, Cassiterite Mining is widespread through Rwanda and numerous other countries, including Nigeria, Malaysia, Bolivia, China, the Democratic Republic of Congo, and Peru, and is the largest source of tin worldwide.^6^ Cassiterite ore (SnO_2_), when refined, has uses in various fields as distinct as semiconductors, electronics, and aeronautics. Methods for collecting Cassiterite entail the digging of pits and veins to access the mineral, and excavation, which often involves hydraulic drilling and blasting. ^7^ There are few if any studies on the occupational hazards of cassiterite mining, though previous retrospective reviews of Chinese tin and tungsten miners showed a high lifetime incidence of silicosis.^8,9^

Additionally, unique risk factors must be acknowledged for those miners operating in Africa. Mining environments are often artisanal or operate without formal regulatory oversight.^10^ Miners in these conditions are at higher potential risk for acquiring silicotuberculosis due to silica dust exposure, cramped working conditions and already higher rates of endemic tuberculosis. Higher rates of HIV can compound the risk for communicable diseases such as TB in the mining environment, ^11^ and silica dust exposure also increases risks of chronic diseases and lung cancer. ^1^

There is paucity of studies to quantify the burden of silicosis in mining communities in Rwanda and East Africa. Current estimates are drawn from screening studies in comparable settings, such as South Africa, Lesotho, and Botswana.^12,13^ In South Africa, the prevalence has been estimated to be approximately five percent amongst gold miners based on survey and chest x-ray screening in an in-service underground cohort. This likely underestimates the fifteen percent prevalence detected on autopsy surveillance possibly due to healthy worker survivor effect, in addition to sub-radiographic silicosis.^14^ Researchers in Botswana studying retired miner cohorts estimated the prevalence of pneumoconiosis to be around 25 percent.^20,21^ Outside of Southern Africa, reports in one East African artisanal mine showed airborne exposures reaching 300-fold greater than the recommended OHSA limit.^16^ Mining for tin, tungsten and other ores has been a feature of the Rwandan economy for decades, with notable under-utilization of administrative and engineering controls, and PPE.^17^

The goal of our study was to better understand the prevalence of silicosis in a representative mining cohort in East Africa, and to complete an analysis of workplace risk.

## Methods

### Study Setting

Kayonza district, located in eastern Rwanda, is a predominantly rural area with 90· 1 % of 344,157 residents residing in rural areas. 10 · 6% of the population has completed secondary or university education. Mean household size is 4· 3 persons. Kayonza has among the lowest population density in Rwanda at 178 inhabitants per square kilometre. 49·8 % of the population is less than 18 years old.^18^

Wolfram Mining and Processing Ltd. (WMP) is a Rwandan company operating mining concessions provided by the Rwandan government and oversees operations at eight semi-industrial mines within the district, with both surface and underground mining activities. Rwandan miners employed by Wolfram Mining and Processing Ltd. that perform underground mining receive training on occupational safety through their employer (based on published company materials).^19^ Rwanda also has a nascent governmental framework for occupational health under the Ministry of Public Service and Labor but continues to work towards adoption of ILO and WHO occupational standards.^20^ All eight mines are in the Rwinkwavu District Hospital (DH) catchment area – a public-operated hospital which supervises eight health centres in southern Kayonza.

### Study Design and Population

We conducted a cross-sectional study of silicosis prevalence among mineworkers in Kayonza district, Rwanda. Eligible participants included all WMP employees at the eight mines in Kayonza district with exposure to underground mining activities. Miners at the time of study were employed in semi-industrial mines that had been upgraded from artisanal in the last 10 years. Per WMP, semi-industrial mines are upgraded from artisanal mines by the inclusion of “adding some facilities in terms of mineral transport, water supply, blasting, jackhammers and the introduction of a small ore processing unit,” Workers were a mix of salaried and “pay for production”. Those recruited directly by our program were active underground miners or employees with previous exposure to underground mining or retired miners who were identified through other miners or our outreach program, who no longer engaged in underground mining. Workers were recruited exclusively through Wolfram. Though the recruited miners work in a semi-industrial workplace, artisanal and small-scale mines are known to operate in Rwanda. ^21^

### Recruitment

Recruitment of mining personnel was anticipated to include all underground miners in Kayonza District who performed regular mining duties through Wolfram Ltd. Initial estimates of underground miners in the district were between 900-1100. Goals of recruitment were to include as many miners within the catchment area as possible in the screening protocol.

### Participant Enrolment and data collection

Between July and August 2022, a total of 1,045 mineworkers at the eight WMP mines in Kayonza were approached by study staff and offered participation in the study. In-person data collection, consisting of a questionnaire and chest radiograph, took place at Rwinkwavu DH between November 1st and December 2nd, 2022. All participants provided written informed consent.

The study team designed an instrument to collect information on demographics; common silicosis symptoms; exposures and risks factors for silicosis, including mine employment history; and past medical history (Appendix). Questions on silicosis risk factors were derived from an existing questionnaire used for silicosis screening.^22^ The instrument was developed in English and translated into Kinyarwanda by the PIH/IMB research team and then reviewed by the Rwandan clinical team to ensure validity for Kinyarwanda speaking patients. Data collectors read questions out loud to the patients and recorded their responses. Questionnaire data were collected and managed using REDCap electronic data capture tools hosted at Partners in Health/Inshuti Mu Buzima.^23^

Analog posterior-anterior chest x-rays were performed by a radiology technician at Rwinkwavu district hospital, then digitized and loaded into a picture archiving and communication system (PACS) for review by the radiology team.

### Key variables

The primary outcome, radiographic silicosis, is a binary variable based on a profusion category of 1/0 or above on chest radiography according to the 2022 International Labor Organization’s (ILO) International Classification of Radiographs of Pneumoconiosis.^24^ Blind review of all chest radiographs was conducted by two independent radiologists—one at University Teaching Hospital of Butare (CHUB) in Rwanda and one at Yale New Haven Hospital—using the ILO classification system. All radiologists were trained in using the ILO Classification. When these radiologists disagreed on a radiographic diagnosis of silicosis, an additional radiologist from Yale New Haven Hospital was used as an arbitrator. The arbitrating third radiologist made the final decision about whether the image was positive for silicosis. Both U.S.-based radiologists were attending-level faculty with specialization in thoracic radiology, and the Rwandan radiologist was head of a medical imaging department at a Rwandan teaching hospital. This arbiter did not review all images, whereas the first two reviewers reviewed all obtained radiographs of sufficient quality. Readers were adherent to ILO guidelines on reading digital chest radiographs in both reading and interpretation.^30^ Radiologists also commented on potential associated diagnoses for actionable feedback from the clinical team. Tuberculosis testing with GeneXpert was obtained on all patients with abnormal chest radiographs denoted by radiology team and who were able to produce the required sputum sample. *“Ubudehe category”* is a socioeconomic status measure using the 2015 Rwandan Government’s four-level categorization of households, ranging from 1 (poorest) to 4 (richest). Unknown ubudehe category was treated as missing data in this analysis.

### Missing data

Patients with missing questionnaire, demographic or radiographic data had those data removed from logistic regression models. All efforts were made to include partially completed questionnaires for patients with acceptable chest X-rays in the analysis. Those chest X-ray films that were uninterpretable due to insufficient quality were removed from the binary variable analysis and excluded from calculations. One positively identified radiographic case of silicosis lacked occupational data and was excluded from the primary analysis but included in inter-rater agreement calculations.

### Statistical Analysis

We described the characteristics of study participants using frequencies and percentages for categorical data and median and interquartile ranges for continuous data. In the bivariate analysis, we used Fisher’s exact and Wilcoxon ranksum tests to measure the association between characteristics of miners and screening positive for silicosis. We fit univariate logistic regression models to assess the association between each covariate and screening positive for silicosis for all variables associated with the outcome at α<0·10 in the bivariate analysis. To adjust for confounding, we also conducted a multivariable logistic regression analysis to investigate factors associated with the outcome. Then, we built a final reduced model using backward stepwise logistic regression in which only variables with a p-value<0· 05 were retained. P-Values were calculated for relevant comparisons. P-values of <0·05 indicates a trend toward statistical significance, using the conventional 95% certainty. Confidence intervals reflect the 95% confidence level. Age and the duration of employment in mining variables were kept in the final model regardless of their p-values. We used Cohen’s Kappa for calculating inter-rater variability between the two primary radiographic raters to account for potential random overlap. We used Wald tests to calculate p-values for each variable in the logistic regression models and all analyses were conducted using Stata v.15·1 (Stata Corp, College Station, TX, USA).

**Figure 1.**
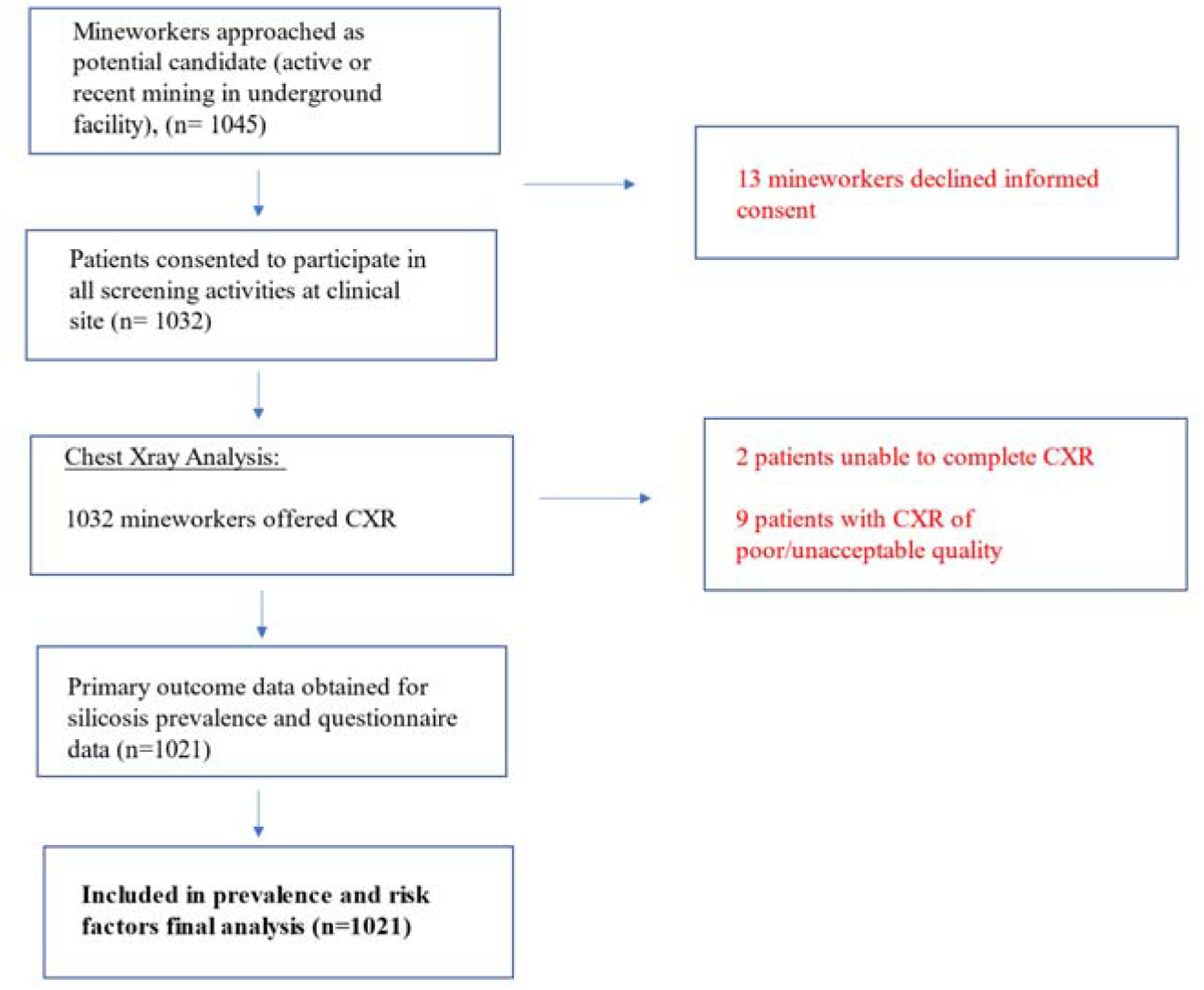
Flow Diagram Demonstrating Recruitment and Analysis.

### Ethics Approval

This study program was approved by Inshuti Mu Buzima Research Committee (IMBRC) and th Rwandan National Ethics Committee (RNEC) 110/RNEC/2022.

#### Funding

This research received no specific grant from an NGO, not-for profit, private, commercial or federal funding source. It was funded internally under the Non-Communicable Diseases budget of Partners In Health-Rwanda otherwise known as Inshuti Mu Buzima.

## Results

### Participants’ characteristics

A total of 1,045 WMP employees were invited to participate in the silicosis case finding program, of which 1,032 (99%) enrolled in the study, with 13 declining informed consent. Two patients who completed informed consent dropped out of the study before collection of data. Data collection was completed for 1,030 for primary outcome analysis – however, 9 participants with poor quality CXR data were excluded from the analysis. Overall, most participants (n=956; 93·3%) were male and the median age was 32 years (IQR 26-40). For those who screened negative for silicosis, the median age was 32 with IQR of [25-38], and those that screened positive, the median age was 41 with IQR of [33-49]. The median duration of working in mining was 7 years (IQR 3-13) (Table 2). For those diagnosed with silicosis, the median time in mining was 15 years (IQR 10-18), with a bottom quartile of <10 years in mining. 43 (4%) reported to have retired from mine work. 20 out of 43 (47%) of these patients screened positive for silicosis, which accounted for 21% of silicosis cases within the cohort.

**Table 1.** Socio-demographic characteristics and health status (N=1030 unless otherwise indicated)

| **Characteristic** | **Total** | **Screened negative for silicosis (N=929)** | **Screened positive for silicosis (N=94)** | **p-value** |
| --- | --- | --- | --- | --- |
| **Socio-demographic characteristics** |  |  |  |  |
| **Gender (N=1023)** |  |  |  |  |
| Male | 956 (93%) | 863 (93%) | 93 (98%) |  |
| Female | 67 (7%) | 66 (7%) | 1 (1%) |  |
| **Age (in years) (N=1023), median [IQR]** | 32 [26-40] | 32 [25-38] | 41 [33-49] | <0.0001 |
| **Marital status (N=1023)** |  |  |  |  |
| Single | 175 (17%) | 173 (19%) | 1 (1%) |  |
| Legally Married | 408 (40%) | 351 (38%) | 57 (61%) |  |
| Cohabiting | 404 (39%) | 369 (40%) | 35 (37%) |  |
| Divorced/Widowed | 37 (4%) | 36 (4%) | 1 (1%) |  |
| **Socio economic status (Ubudehe) (N=1008)** |  |  |  |  |
| Category 1 | 96 (9%) | 89 (10%) | 7 (8%) |  |
| Category 2 | 481 (48%) | 488 (49%) | 33 (35%) |  |
| Category 3 | 431 (43%) | 377 (41%) | 54 (57%) |  |
| **Health status** |  |  |  |  |
| **BMI, median [IQR]** | 21.1 [19.8-22.6] | 21.2 [19.9-22.7] | 20.2 [18.8-21.3] | <0.0001 |
| **Self-reported History** |  |  |  |  |
| HIV infection | 29 (3%) | 26 (3%) | 3 (3%) | 0·57 |
| Exposed to tobacco (N=1,007) | 328 (33%) | 268 (29%) | 60 (63%) | <0·0001 |

**Table 2.** Occupational characteristics of Screened Miners.

| Characteristic | Total | Screened negative for silicosis (N = 929) | Screened positive for silicosis (N = 94) | p-value |
| --- | --- | --- | --- | --- |
| **Exposure to silica dust** |  |  |  |  |
| Duration of working in mining (in years) (N=1,020), median [IQR] | 7 [3-13] | 6 [3-12] | 15 [10-18] | <0.0001 |
| Currently working in mining (N=1,027) | 984 (96%) | 909 (98%) | 75 (79%) | <0.0001 |
| Ever worked in construction (N=1,007) | 197 (20%) | 187 (20%) | 10 (11%) | 0.02 |
| Not currently working in mining (Retired) (N = 1,027) | 43 (4%) | 23 (2%) | 20 (21%) | <0.0001 |
| **Duty stations** |  |  |  |  |
| Ever worked in drilling station (N=1,023) | 460 (45%) | 392 (42%) | 68 (72%) | <0.0001 |
| Ever worked in excavation station (N=1,023) | 825 (81%) | 737 (79%) | 88 (94%) | <0.0001 |
| Ever worked in blasting station (N=1,023) | 340 (33%) | 291 (31%) | 49 (52%) | <0.0001 |
| Ever worked in solid disposal station (N=1,023) | 569 (56%) | 528 (57%) | 41 (44%) | 0.02 |
| Ever worked in washing harvest (N=1,023) | 539 (53%) | 493 (53%) | 46 (49%) | 0.45 |
| Working in multiple duty stations (N=1,023) | 710 (69%) | 635 (68%) | 75 (80%) | 0.03 |
| **Protection against silica hazards** |  |  |  |  |
| Availability of facemask (N=1,022) | 86 (8%) | 74 (8%) | 12 (13%) | 0.12 |
| **Use water while drilling (N=1,023)** |  |  |  |  |
| No | 62 (6%) | 49 (5%) | 13 (14%) |  |
| Yes | 920 (90%) | 840 (90%) | 80 (85%) |  |
| NA | 41 (4%) | 40 (4%) | 1 (1%) |  |

### Prevalence of silicosis and associated factors

Among 1,021 participants with data on silicosis status and occupation status, the prevalence of radiologic silicosis was 9% (Table 3). In the final adjusted logistic regression model, increased odds of silicosis were associated with having retired from the mine work before the screening date (aOR 8·88; 95% CI 3·87-20·38), self-reported medical history of tuberculosis diagnosis (aOR 4·15; 95% CI 1·42-12·11), working in blasting station (aOR 3·30; 95% CI 1· 68-6·45), excavation station (aOR 2· 77; 95% CI 1·09-7· 04), drilling station (aOR 2·51; 95% CI 1·34-4·70), being exposed to tobacco (aOR 1·92; 95% CI 1·14-3·24), increased time spent working in mining (aOR 1·05; 95% CI 1·01-1·09), and age (aOR 1·05; 95% CI 1·02-1·09). Lower odds of silicosis were observed among participants working in solid disposal station (aOR 0·25; 95% CI 0·14-0·47) and inversely associated with BMI (aOR 0·86; 95% CI 0·75-0·97).

**Table 3:** Assessing the factors associated with screening positive for silicosis.

| Variables | Univariate analysis of the association between each covariate and screening positive for silicosis |  | Multivariable analysis of the association between covariates and screening positive for silicosis |  |  |  |
| --- | --- | --- | --- | --- | --- | --- |
|  |  |  | Full model<sup>2</sup> (n=974) |  | Reduced model<sup>3</sup> (n=989) |  |
|  | <sup>1</sup>OR [95% CI] | p-value | aOR<sup>4</sup> [95% CI<sup>5</sup>] | p-value | aOR<sup>4</sup> [95% CI<sup>5</sup>] | p-value |
| **Socio-demographic characteristics** |  |  |  |  |  |  |
| **Gender** |  | 0.06 |  | 0.71 |  |  |
| Male | ref |  | ref |  |  |  |
| Female | 0.14 [0.02-1.04] |  | 0.65 [0.07-5.91] |  |  |  |
| **Age (years)** | 1.08 [1.06-1.10] | <0.0001 | 1.05 [1.01-1.08] | 0.01 | 1.05 [1.02-1.09] | 0.001 |
| **Ubudehe category** |  | 0.03 |  | 0.42 |  |  |
| Category 1 | ref |  | ref |  |  |  |
| Category 2 | 0.95 [0.41-2.21] |  | 1.31 [0.45-3.84] |  |  |  |
| Category 3 | 1.80 [0.79-4.10] |  | 1.74 [0.61-4.96] |  |  |  |
| Unknown | 0.61 [0.07-5.19] |  | - |  |  |  |
| **Health status** |  |  |  |  |  |  |
| **BMI (kg/m<sup>2</sup>)** | 0.76 [0.68-0.85] | <0.0001 | 0.87 [0.76-0.99] | 0.04 | 0.86 [0.75-0.97] | 0.02 |
| **Self-reported medical history** |  |  |  |  |  |  |
| **Tuberculosis** |  | 0.003 |  | 0.02 |  | 0.01 |
| No | ref |  | ref |  | ref |  |
| Yes | 3.86 [1.58-9.45] |  | 3.90 [1.20-12.62] |  | 4.15 [1.42-12.11] |  |
| **Exposed to tobacco** |  | <0.0001 |  | 0.01 |  | 0.01 |
| No | ref |  | ref |  | ref |  |
| Yes | 4.08 [2.62-6.35] |  | 2.14 [1.24-3.70] |  | 1.92 [1.14-3.24] |  |
| **Occupational characteristics** |  |  |  |  |  |  |
| **Duration of working in mining (in years)** | 1.12 [1.09-1.15] | <0.0001 | 1.05 [1.01-1.10] | 0.02 | 1.05 [1.01-1.09] | 0.02 |
| **Currently employed in mining** |  | <0.0001 |  | <0.0001 |  | <0.0001 |
| No | 11.07 [5.78-21.21] |  | 8.76 [3.45-22.21] |  | 8.88 [3.87-20.38] |  |
| Yes | ref |  | ref |  | ref |  |
| **Ever worked in construction** |  | 0.03 |  | 0.38 |  |  |
| No | ref |  | ref |  |  |  |
| Yes | 0.47 [0.24-0.92] |  | 0.69 [0.31-1.56] |  |  |  |
| **Duty stations** |  |  |  |  |  |  |
| **Ever worked in drilling station** |  | <0.0001 |  | 0.02 |  | 0.004 |
| No | ref |  | ref |  | ref |  |
| Yes | 3.59 [2.24-5.75] |  | 2.70 [1.19-6.14] |  | 2.51 [1.34-4.70] |  |
| **Ever worked in excavation station** |  | 0.002 |  | 0.09 |  | 0.03 |
| No | ref |  | ref |  | ref |  |
| Yes | 3.79 [1.63-8.81] |  | 2.33 [0.86-6.31] |  | 2.77 [1.09-7.04] |  |
| **Ever worked in blasting station** |  | 0.0001 |  | 0.001 |  | 0.001 |
| No | ref |  | ref |  | ref |  |
| Yes | 2.37 [1.54-3.64] |  | 3.48 [1.72-7.02] |  | 3.30 [1.68-6.45] |  |
| **Ever worked in solid disposal station** |  | 0.01 |  | 0.0002 |  | <0.0001 |
| No | ref |  | ref |  | ref |  |
| Yes | 0.57 [0.37-0.88] |  | 0.29 [0.15-0.56] |  | 0.25 [0.14-0.47] |  |
| **Worked in multiple duty stations** |  | 0.03 |  | 0.34 |  |  |
| No | ref |  | ref |  |  |  |
| Yes | 1.82 [1.08-3.07] |  | 0.62 [0.24-1.66] |  |  |  |
| **Protection against silica hazards** |  |  |  |  |  |  |
| **Use water while drilling** |  | 0.003 |  | 0.75 |  |  |
| No | ref |  | ref |  |  |  |
| Yes | 0.35 [0.18-0.67] |  | 0.88 [0.34-2.28] |  |  |  |
|  | 0.09 [0.01-0.74] |  | 0.40 [0.04-4.30] |  |  |  |

<sup>1</sup>Unadjusted odds ratio from univariate models of the association between each covariate and the outcome
<sup>2</sup>The full model included all variables associated with the outcome at $\alpha < 0.10$ in the bivariate analysis – however, we excluded from the model the self-reported medical history of silicosis diagnosis, as missing data for this variable was at 18%.
<sup>3</sup>The reduced model which retained variables with a p-value < 0.05
<sup>4</sup>**aOR**, adjusted odds ratio
<sup>5</sup>**CI**, confidence interval
**n**, number of observations included in the model

### Radiographic profile of silicosis

Two trained radiologists provided analysis of all radiographs of good or acceptable quality. 1,011 radiographs were deemed of “good” and 10 radiographs of “acceptable” quality were included in the analysis. Of the 1,021 radiographs reviewed, Radiologist #1 interpreted 93/1021 (9·1%) and Radiologist #2 interpreted 86/1021 (8 · 4%) as radiographically positive for silicosis based on ILO criteria. In accordance with ILO guidelines, radiologists also rated positive films with large opacities with *ABC* ratings and small opacities with *pqr* (small, regular) and *stu* (small, irregular) ratings (table 4). 61/93 (65·5%) and 50/86 (58·1%) of our silicosis patients ILO rated perfusion scores of 2/1 or above based on our first and second rater respectively. 15·1% (14/93) and 14% (10/86) of radiographs showed “large” opacification respectively. Our first rater noted 53 films with irregular (*stu)* small opacification and our second rater noted 36 films with irregular opacities.

**Table 4:** Clinical and radiological profile of silicosis among miners.

|  | Radiologic silicosis (Reviewer 1) |  |  | Radiologic silicosis (Reviewer 2) |  |  |
| --- | --- | --- | --- | --- | --- | --- |
|  | No silicosis suspected, n (%) | Silicosis suspected, n (%) | P-value | No silicosis suspected, n (%) | Silicosis suspected, n (%) | P-value |
| **Symptoms** |  |  |  |  |  |  |
| Persistence of dry cough in the past 2 week | N=928<br>153 (16.5) | N=93<br>29 (31.2) | 0.001 | N=935<br>154 (16.5) | N=86<br>29 (33.7) | <0.001 |
| Shortness of breath in the past 2 week | N=906<br>257 (28.4) | N=93<br>54 (58.1) | <0.001 | N=912<br>259 (28.4) | N=86<br>54 (62.8) | <0.001 |
| Weakness in the past 2 weeks | N=903<br>382 (42.3) | N=92<br>60 (65.2) | <0.001 | N=909<br>382 (42.0) | N=85<br>58 (68.2) | <0.001 |
| Chest pain in the past 2 weeks | N=918<br>393 (42.8) | N=93<br>61 (65.6) | <0.001 | N=926<br>396 (42.8) | N=85<br>59 (69.4) | <0.001 |
| Loss of appetite in the past 2 weeks | N=918<br>231 (25.2) | N=93<br>48 (51.6) | <0.001 | N=926<br>235 (25.4) | N=85<br>45 (52.9) | <0.001 |
| Wasting and weight loss in past 2 weeks | N=918<br>393 (42.8) | N=93<br>48 (51.6) | 0.124 | N=926<br>398 (43.0) | N=85<br>46 (54.1) | 0.052 |
| **ILO categories** |  |  |  |  |  |  |
| *Received ILO rating from interpreter* | - | N=93<br>79 (84.9) |  | - | N=86<br>76 (88.4) |  |
| **ILO grade 0/0** | - | N=93<br>0 |  | - | N=86<br>0 |  |
| **ILO grade 0/1** | - | N=93<br>7 (7.5) |  | - | N=86<br>11 (12.9) |  |
| **ILO grade 1/1** | - | N=93<br>3 (3.2) |  | - | N=86<br>8 (9.3) |  |
| **ILO grade 1/2** | - | N=93<br>0 |  | - | N=86<br>7 (8.1) |  |
| **ILO grade 2/+** | - | N=93<br>50 (53.7) |  | - | N=86<br>40 (46.5) |  |
| **ILO grade 3/+** | - | N=93<br>11 (11.8) |  | - | N=86<br>10 (11.6) |  |
| **CXR abnormalities** |  |  |  |  |  |  |
| **Size of opacity observed** |  |  | - |  |  | - |
| Both large and small opacity | - | 19 (20.4) |  | - | 24 (27.9) |  |
| Large (>1cm) | - | 14 (15.1) |  | - | 12 (14.0) |  |
| Category A | - | 10 |  | - | 9 |  |
| Category B | - | 13 |  | - | 16 |  |
| Category C | - | 10 |  | - | 11 |  |
| Small (=<1cm) | - | 78 (83.9) |  | - | 70 (81.4) |  |
| Category p/- | - | 8 |  | - | 25 |  |
| Category q/- | - | 10 |  | - | 9 |  |
| Category r/- | - | 3 |  | - | 0 |  |
| Category s/- | - | 11 |  | - | 2 |  |
| Category t/- | - | 32 |  | - | 23 |  |
| Category u/- | - | 9 |  | - | 11 |  |
| Mixed Category | - | 5 |  | - | 0 |  |
| Total Agreement |  | 80/95 |  |  |  |  |
| Cohen's inter-rater<br>variability<sup>1</sup> for ILO >1/0 |  | **k=0.837** |  |  |  |  |

<sup>1</sup>Cohen's test defined as $k = (\mathbf{po} - \mathbf{pe}) / (1 - \mathbf{pe})$ where **po**: Relative observed agreement among raters and **pe**: Hypothetical probability

Total number of agreements between the two radiologists was 80 out of 95 total positive ratings, with an inter-rater reliability of 0·842. Using Cohen’s Kappa test to account for random overlap, **k**=0·837. All mine workers with a diagnosis of silicosis were brought back to the NCD clinic within 6 months of the completion of data analysis for disclosure of diagnosis. A follow up plan based on clinical severity of silicosis was developed for the cohort. ^25^

## Discussion

This paper is the first to measure the silicosis prevalence and associated risk factors among cassiterite miners in Rwanda, and to our knowledge, East Africa. We sought to understand further the occupationally mediated risk factors leading to the high prevalence of silicosis in our mining cohort. Our findings suggest that certain work practices and a relatively dusty environment likely led to increased cumulative inhalation of silica dust in this cohort of cassiterite miners, including blasting and drilling.^26^ We also noted the established risk factors of tuberculosis and smoking contributing to silicosis in our cohort. Studies have noted a possible synergistic effect of lung damage from smoking and silica inhalation leading to silicosis.^27^ Additionally, our survey of personal protective equipment revealed that facemasks were only reported available for 8% of workers. These facemasks were not occupationally rated and primarily made of cloth material. These results suggest further analysis of mining work environment to understand and quantify individual silica exposure is warranted.

In this working age population of miners in rural Rwanda, radiographic prevalence of silicosis approaches 10 percent. Case finding within a working age population offers the distinct opportunity to recruit patients into surveillance programs and clinical care but is likely to have lower positivity due to the lower cumulative exposure to silica dust and lack of latency period compared to studies evaluating retired miners, autopsy studies, or retrospective studies.^12,14^ Additionally, certain finding in our cohort appear noteworthy. 61/93 (65 · 5%) and 50/86 (58 · 1%) of our silicosis patient presented with ILO rated profusion scores of 2/1 and above, indicating a preponderance of higher profusion scores that typically correlate with more advanced disease. This could be a function of the inclusion of retired miners (a group that had radiographic silicosis at a rate of 47% in this cohort), but this cannot account for all these ratings. Many patients reported symptoms of shortness of breath, weakness and cough within our cohort, which was statistically higher within our silicosis patients. Despite this, only 5 patients tested positive for active tuberculosis among our cohort in sputum testing.^25^ Possibly these findings could be related to a preponderance of more radiographically advanced disease (ILO 2/1 and above) and large opacities (15·1%/14%).

Though Cassiterite itself does not contain silica dust, it often co-occurs in quartz veins due to predilection to form in similar geologic environments, with quartz serving as a host mineral for cassiterite frequently in Rwandan deposits.^28^ Jobs that involve disruption of quartz (such as the drillers and blasters) are likely exposed to high levels of aerosolized silica due to the disruption of quartz, which is known to be associated with silicosis.^29^

Studies suggest that one death from silicosis only estimates 4-8% of cases occurring in the community, and thus silicosis represents a unique disease where clinical diagnosis is often too late to affect the natural history of disease.^30^ Upstream interventions are necessary for disease prevention. Thus, the emphasis of our study was to identify risk factors in mining practice and to identify risk factors that were amenable to intervention in clinical practice.^15,16^

Today, pneumoconiosis is seeing a resurgence in mining communities around the globe, including those in the United States.^31^ Miners in LMICs have additional barriers to accessing medical care due to structural and logistical barriers which can complicate the establishment of a comprehensive silicosis management program. ^32^ Due to the heterogeneity of environments in mining communities, a uniform approach to silicosis case finding has not been agreed upon by consensus and should be guided by availability of local resources.^33^

The Global Burden of Disease study report showed only 8 incident cases of silicosis reported in 2017.^4^ This is despite previous subjective evidence suggesting the problem may be more widespread.^17^ Our work discovered 95 cases of silicosis in a high-risk cohort in one district in the Eastern Province of Rwanda over a 1-month period. We hope this data adds evidence that proactive screening needs to be done for a more accurate reflection of disease prevalence in Rwanda and countries with developing mining sectors.

### Limitations

There may be reservation by employers, employees, and governments in engaging in occupational health case finding and surveillance that may limit the accuracy of these findings. However, with collaboration from local mining companies and the mining community, we were able to recruit a large group of miners into our study. It is the ongoing responsibility of employers, governments, and NGOs to continue to draw attention to this issue and provide safe and adequate working conditions for miners.^34^

There may be some doubt as to the definitive diagnosis of silicosis in our patient population. Similar pneumoconiosis, such as Stannosis or mixed-dust pneumoconiosis, could cause a similar radiographic appearance and during this study we were unable to offer more definitive diagnostic studies such as high-resolution CT scanning or lung biopsy.^35^ However due to characteristic upper and mid-lobe predominance of disease on radiographs and high inter-rater reliability, as well as characteristics of the mining work of these patients (hard rock mining in quartz-rich veins), Silicosis is the favoured diagnosis. There is continued debate as to the ideal screening imaging for silicosis, with some groups recommending Chest CT. Chest X-ray proved most viable logistically for our cohort, though it may underestimate overall prevalence. ^33^

We recognize that mining practices are local and various risks that contribute to a high incidence of silicosis among cassiterite miners in Rwanda may not be present in other Cassiterite mines, though the burden remains on employers to guarantee the safety of their workers. However, similar techniques are employed to harvest cassiterite and thus these data can serve as a reasonable entry into occupational risk regarding cassiterite mining in LMICs.^7^

Finally, our group agreed to provide clinical assistance to Wolfram Inc. for the purposes of silicosis screening for this protocol. We are currently in discussions to perform additional workplace hazard analysis in Wolfram mines. In 2001, the American Conference of Industrial Hygienists adopted a threshold limit value of 0·05 mg/m^3^ for respirable crystalline silica daily, but while there is no known safe respirable concentration of RCS to prevent disease, efforts to maintain lower exposure levels are imperative to prevent incidence and progression of disease.^1^

We also acknowledge the limitation of cross-sectional studies in proving definitive correlations between exposures and disease states. Our group continues to generate longitudinal data in this cohort.

## Conclusions

Our results suggest that the rate of silicosis in mining communities in East Africa are high. Our methodology differed from other screening studies in the region but showed a roughly two-fold greater risk of silicosis than South African gold miners and Chinese tin miners, but lower than tanzanite miners in Tanzania.^36^ We additionally provide evidence that Cassiterite mining employs high-risk techniques that are significantly associated with the development of silicosis in a cross-sectional mining cohort, possibly related to the naturally occurring co-mineralization of cassiterite with quartz. Additional work still needs to be completed. Further characterization of the mining environment with support from local mining companies will better allow us to tailor interventions to the unique mining environments of East Africa. Comprehensive national and international regulations providing for safe working environments for miners in high-risk industries are imperative for control of silica-related disease, and efforts to adopt these conventions globally is imperative for elimination of silicosis. We hope this evidence suggests that physicians and healthcare organizations in LMICs can meet the needs of these populations and collaborate with mining industry to develop effective systems to address silicosis.

## Author Contributions

The authors confirm contribution to the paper as follows and all authors fulfil all four ICJME criteria. Individual contributions are as follows: study conception and design: IK,PB,SH,VKC, DM, SD,NA,WD, PS Data collection: AN,RT,SD, PM, JNR Analysis and interpretation: AN,AN,SD,SH, MM, LT,TE, VKC,FK, PS, JM, MN, TE, LT, JNR. Draft manuscript preparation: SD,SH. Draft manuscript critical review: all authors. All authors reviewed the results and approved the final version of the manuscripts.

## Data Sharing Statement

The authors acknowledge that data collected for this study, including participant data can be made available to others per Rwandan law. Data that is made available will be deidentified and includes questionnaire data, radiography data and other demographic data. Study protocol can be viewed on clinicaltrials.gov at NCT05538299. Informed consent form can be provided on request. Per Rwandan patient confidentiality law, patient data cannot be made publicly available. For parties interested in obtaining study data, they will need to submit a data sharing/access agreement to Inshuti Mu Buzima which will need to be approved by the Rwandan Data Security Office. Data can be made available by request after the publication date. Requests can be initiated by communication with the corresponding author

